# Exploring new EEG-parameters in electroconvulsive therapy

**DOI:** 10.1101/2021.11.03.21265830

**Authors:** J Schreiner, U Kessler, KJ Oedegaard, KA Mardal, L Oltedal

**Author notes:** **Corresponding Author:** Jakob Schreiner^a, b^.

## Abstract

**Background:** Electroconvulsive therapy (ECT) is an effective treatment against severe depressive episodes, which has been shown to induce volume changes in the hippocampus. The power spectrum of the electroencephalogram (EEG) follows a characteristic power-law relation but its utility as a metric of ECT-induced seizures has not been explored.

**Objective:** This study aims to evaluate a novel metric based on the power spectrum of the EEG recordings from ECT-induced seizures; its association to volume changes in the hippocampus following ECT and improvement in depression rating scores.

**Methods:** Depressed patients treated with ECT underwent brain MRI before- and after treatment and the EEG from each seizure was recorded (N=29). Hippocampal volume changes and EEG parameters were recorded in addition to clinician-rated and self-reported measures of depressive symptoms. The slope of the power-law in the power spectral density of the EEG was calculated. Multivariate linear models relating seizure parameters to volume change or clinical outcome was systematically and successively simplified. The best models were selected according to Akaike’s information criterion.

**Results:** The slope of the power-law was steeper in the right than the left hemisphere *(p* < 0.001). EEG measures were included in the best models of volume change for both hippocampi as well as in the models explaining clinical outcome (*p* = 0.014, *p* = 0.004).

**Conclusions:** A novel EEG measures was explored and contributed in models explaining the variation in volume change in the hippocampus and in clinical outcome following ECT.

## Introduction

Electroconvulsive therapy (ECT) is the most effective treatment used against severe depressive episodes [1]. The procedure involves general anesthesia and treatment electrodes placed on the skull to deliver electrical stimuli. Monitoring electrodes record the resulting seizures as an electroencephalogram (EEG). Normally, this procedure is repeated twice or thrice weekly for a total of about 8 -12 sessions.

Objective, quantitative inter-individual measures of ECT-induced effects on the brain can be provided by both Magnetic Resonance Imaging (MRI) and EEG. MRI enables detailed spatial analysis of brain morphology but has a poor resolution in time. EEG on the other hand, describes the temporal course of a seizure but is typically limited to a few localized probes. While EEG has been widely used in clinical practice to monitor ECT-induced seizure activity since at least the 1980s [2], MRI has been used for ECT research over the last three decades [3]. There is, however, a lack of studies combining both measures.

There is a longstanding line of research with the aim of optimizing the stimulus parameters to reduce the cognitive side effects while maintaining the antidepressant efficacy [4]. Several EEG parameters have been used to monitor the adequacy of the seizure. Seizure duration alone is not sufficient as a marker of effective treatment [5-7]. Nobler et al. [8] proposed a mixture of qualitative and quantitative features of the ictal EEG that could guide electrical stimulus dosing while Krystal et al. [9] demonstrated the use of computer derived ictal EEG measures to distinguish between electrode placement and stimulus dosage. Nobler et al. [10] found a correlation between ictal EEG parameters and treatment outcomes, and speculated that algorithms based on EEG parameters might serve ECT practitioners in adjusting the dosage to account for increased seizure thresholds. Although monitoring of seizure quality has been recommended across the course of treatment [11], the general guidelines for monitoring ECT are still vague and there is considerable inter-individual variation [12].

EEGs are commonly analyzed in terms of their frequency components grouped into delta (0.7 – 3.5 Hz), theta (3.5 - 8 Hz), alpha (8 - 13 Hz), beta (13 - 25 Hz), and gamma (25 -> Hz), both under normal function and during seizures. The spectral power in these waves characteristically drops several orders of magnitude from low to high frequencies [13] and can be characterized in terms of a power-law *p* ∼ *f*^*a*^, where *f* is the frequency and a is negative. A clear benefit of the power-law description is that it reduces the complexity of the EEG across all (or a range of) frequencies to one number that describes the relationship between the energy of the different waves. Under normal brain activity Milstein et al. [14] found a = -2 for frequencies less than 400 Hz and Miller et al. [15] found a = -2 for frequencies less than 75 Hz and a = -4 for frequencies between 80 and 500 Hz. In a simulation study, Pettersen et al. [16] found a to be between -0.5 and -3. The use of the power-law description has to our knowledge not been investigated in the context of ECT, but there are differences in the relative power in the frequency bands during ECT-induced seizures [17]. Both the differences between the waves and how they change throughout the seizure are captured in the power-law relationship.

Morphological changes in the brain following ECT, and to which extent brain regions express different responses, have been the focus of several studies using magnetic resonance imaging. Volumetric changes in the gray matter following ECT were first observed in the hippocampus [18, 19], although these changes are not only restricted to the medial temporal lobe [3]. A recent study suggested that some volume change in the grey matter is apparent 2 hours after the first ECT [20].

The volumetric changes are known to correlate with the number of treatments given in an ECT series, to depend on electrode positions [19] and to correlate with the local electrical field [21, 22]. However, Takamiya et al. found the volume expansion in the medial temporal lobe correlated with the cumulative seizure duration, and not the simulated electrical field [22]. Moreover, the behavioral significance of gray matter volume changes has not been established [23].

In this study, we explore whether summary metrics including the power-law behavior of the ictal EEG can explain the volume changes in the hippocampus. Furthermore, we investigate whether including both the volume change in the hippocampus and EEG metrics, using EEG leads from both hemispheres, can inform predictions of clinical outcomes.

## Material and Methods

### Patient Characteristics

Data from the current project stem from a study assessing ECT-related brain changes as measured by MRI. This study protocol is previously described in detail elsewhere [24]. MRI before and after the completion of the ECT treatment was available from 29 patients (14 female, age 46.4 ± 15.9). All patients participating in the study received ECT with right unilateral (RUL) electrode placement, except one who switched to bilateral electrode placement for the last four treatments. The number of treatments varied from 3 to 20 (9.9 ± 4.3). The initial charge was based on age and sex, and later adjusted, see [24]. All participants provided written and informed consent and the study was approved by the Regional Committee for Medical and Health Research Ethics, REC Southeast Norway (2013/1032).

### Image and EEG Acquisition

EEGs were recorded by the Thymatron IV [25] machine, which also was used to administer the electrical stimulus. The operator was instructed to let the upper and lower EEG electrodes to be placed over the right and left hemispheres respectively. In the statistical analysis in this paper, we used the measures time to peak coherence (TPC) and maximum sustained coherence (MSC), which were computed and automatically provided by the Thymatron IV. Two Thymatron IV machines were used in this study. TPC, MSC and PSI are the only EEG parameters computed by both machines. However, PSI was computed by different algorithms in the two Thymatron IV machines. Therefore, postictal suppression (PS) was rated by the operator on a scale from 1 to 3. Compared to the reference EEG taken before the treatment, 1 means an almost completely flat EEG, 2 means a visibly lower EEG amplitudes, and 3 means no discernible differences. In addition to these metrics, we also used the cumulative seizure duration over all the treatments for each patient, as also done by Takamiya et al. [22]. The seizure duration was determined by the operator. Furthermore, we computed the spectral energies in the delta, theta, alpha, and beta frequency bands; see e.g. [10, 26, 27]. The energies in these bands were normalized by the total energy computed over the whole frequency spectrum. Finally, we computed the power-law exponent in the power spectral density (PSD) of the EEG between 4 and 75 Hz, following the power-law documented by Miller et al. [15]. All parameters were averaged across all treatments each patient received. The power law and the energies within the four frequency bands were computed and averaged for the left and right hemispheres separately [27]. In addition to the ictal EEG features, age, sex, and number of treatments and seizure duration was included in the statistical models.

### Imaging

Details of the 3 Tesla Magnetic Resonance Imaging acquisitions and processing have been described elsewhere [20, 24], and the same image pre-processing steps were adopted here. The brain MRIs were next processed using FreeSurfer version 7.1.1 (https://surfer.nmr.mgh.harvard.edu), which provides automated segmentation of subcortical deep gray matter structures [28, 29]. The longitudinal stream [30] was applied to compute the volume changes in the hippocampus.

### Depression Rating Scale

To assess the clinical outcome of ECT for depression, two measures capturing characteristic attitudes and symptoms of depression were used; the self-reported Beck Depression Inventory (BDI) [31] and the clinician-rated Montgomery and Åsberg Depression Rating Scale (MADRS) [32].

The EEG strips generated by the Thymatron IV were digitized as described in Appendix A. The PSD is estimated using Welch method [33] implemented in *scipy* 1.6.2 [34].

### Statistical Methods

We created a series of linear models explaining the volume change in the left and right hippocampus separately in terms of the EEG parameters, age, sex, and ECT dosage. A general model with 14 regressors: number of sessions, cumulative charge, cumulative seizure duration, Age, sex, PS, MSC, TPC, least squares (LSQ) and least upper (LU) approximations to the slope of the power-law, and the fraction of the energy in the delta, theta, alpha and beta bands. The model was successively simplified through a backward elimination procedure, similar to Perera et al. [27]. In each iteration of the procedure, the regressor in the linear model having the *t*-value with the lowest magnitude was removed. Akaike’s information criterion (AIC) was used to select the best model among the successively simplified models. We verified that the best model also minimizes the *consistent* AIC (cAIC). cAIC is defined as the average of AIC and BIC (Bayesian information criterion), and places a greater penalty on the number of regressors [35]. The linear models were created using the *lm* function from the software package R version 4.0.2 [36]. The same model simplification procedure was adopted for exploring models relating volume change in the hippocampus and the EEG parameters to change in MADRS and BDI. Starting with a general model with 15 regressors: volume change in the left hippocampus and the right hippocampus, number of ECT sessions, cumulative charge, cumulative seizure duration, age, sex, MSC, and TPC. In the interest of limiting the number of regressors due to limited sample size, and including information from both EEG leads as well as the volume change in the hippocampus we excluded several parameters that we used in the models of volume change in the hippocampus. We included only LSQ, as it is more sensitive to the PSD of the whole frequency spectrum than LU. Furthermore, we only used the delta and beta bands from the left hemisphere and alpha and beta bands from the right hemisphere, as these were the frequency bands that performed the best in the models of volume change in the left and right hippocampus.

## Results

The slope of the power-law, *a*, was estimated with LSQ in the frequency interval 4 Hz to 75 Hz, shown in Figure 1a. The energy drops off four orders of magnitude in this frequency interval. Noting the consistent presence of two peaks in the delta and beta bands (Figure 1b), we also computed a, using LU, such that the power-law is the least upper bound of the PSD. LSQ and LU were highly correlated and produce similar distributions but with different means, see Figure 2a. The Pearson correlation coefficient between LSQ and LU was *r* = 0.85 in the left and *r* = 0.99 in the right hemisphere. The LSQ measure was different between the left (−3.8 ± 0.4) and right (−4.1 ± 0.2) hemispheres, *t*(28) = 5.17, *p* < 0.001. The LU measure is also different between the left (−3.6 ±0.4) and right (−4.1 ± 0.2) hemispheres, *t*(28) = 7.08, *p* < 0.001. The fraction of the total energy in each of the frequency bands was similar in the left and right hemispheres except in the theta ban, *t*(28) = -2.70, *p* = 0.011, see Figure 2b. The majority of the energy was found in the lower frequencies.

**Figure 1:**
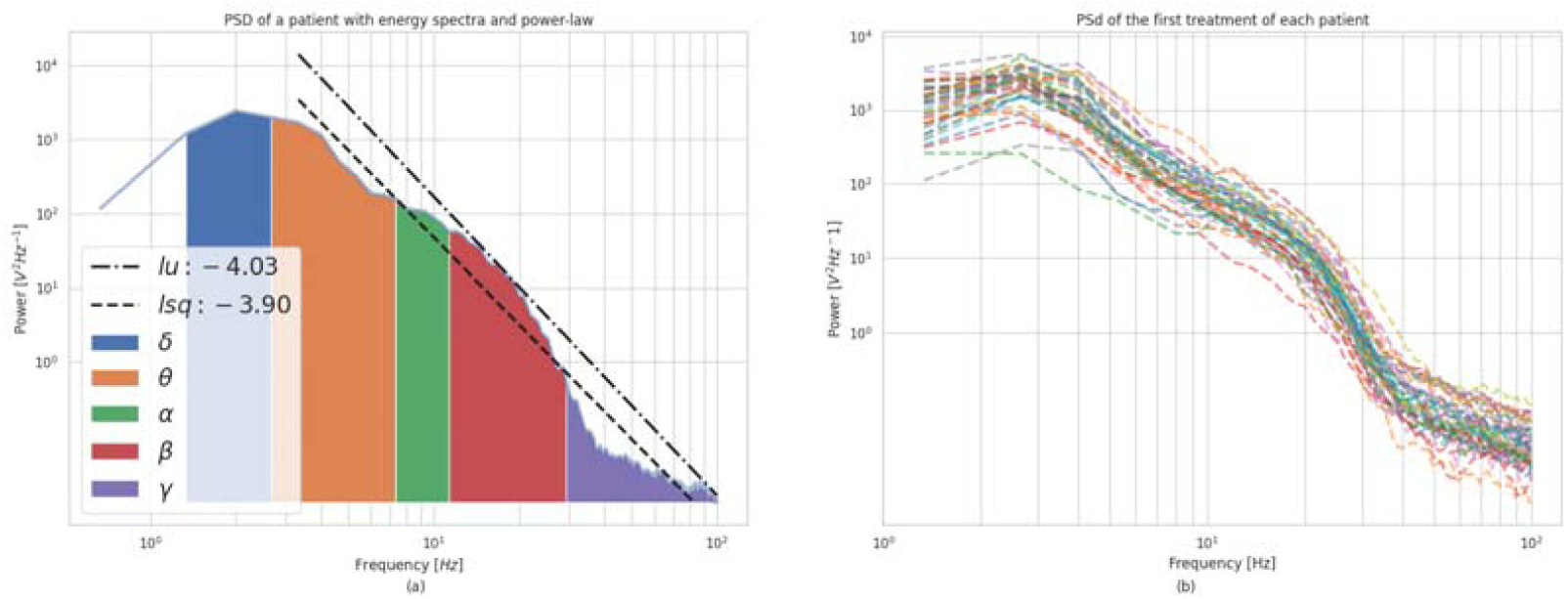
Power-law: (a) The power spectral density (PSD) of a single seizure with two lines illustrating power-law with a least squares (LSQ) and least upper (LU). The area under the curve is colored delta (0.7 – 3.5 Hz), theta (3.5 – 8 Hz), alpha (8 – 13 Hz), beta (13 – 25 Hz), gamma (> 25 Hz) waves. The area is proportional to the energy associated with each of the waves. (b) The PSD for the first seizure for each patient. Both figures are from an electroencephalogram EEG of the left hemisphere.

**Figure 2:**
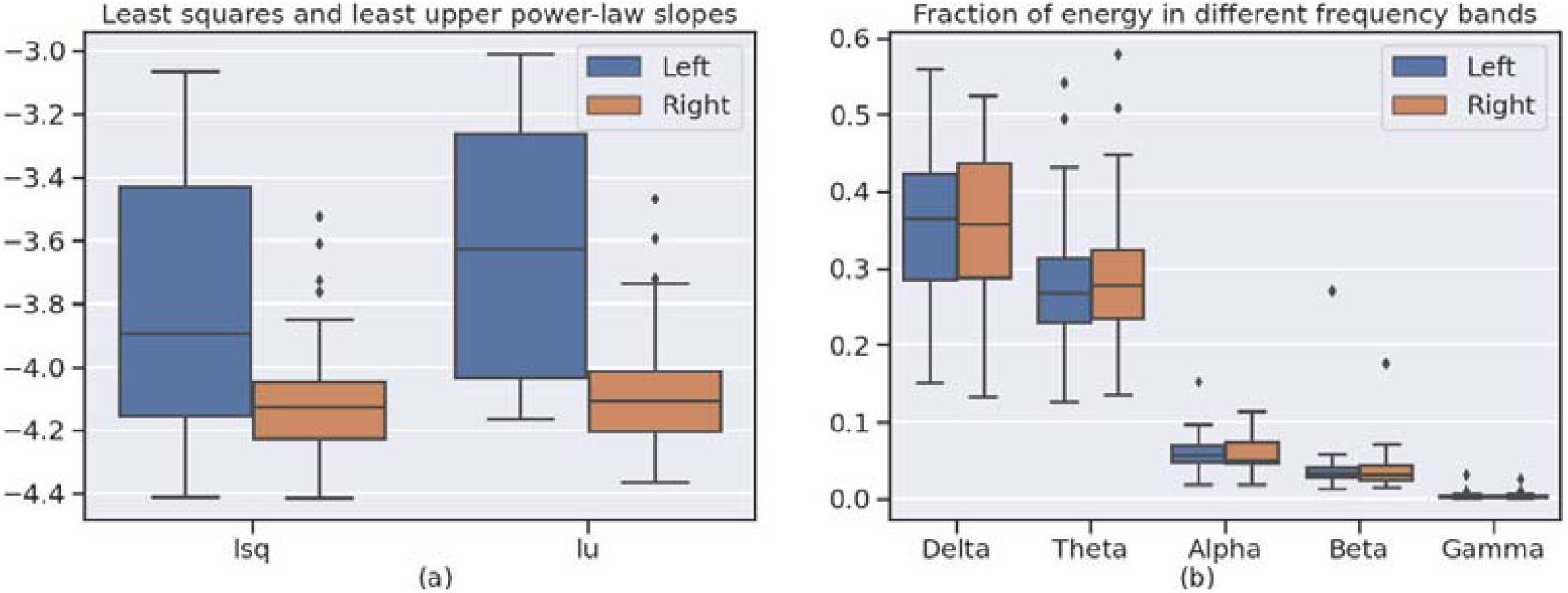
Power-law and spectral energy: Distributions of the slope of the power laws (a), and the energies in different frequency bands (b) for the left and right hemispheres. Volume change in left and right hippocampus from pre to post ECT is shown in Figure 3a. The mean ± standard deviation change in the left hippocampus was 76.29 ± 84.08 mm^3^ and 140.63 ± 95.52 mm^3^ in the right hippocampus. The change in MADRS and BDI is shown in Figure 3b. MADRS and BDI were 33.1 ± 5.8 and 34.6 ± 12.2 before the first treatment and 15.7 ± 9.1 and 19.2 ± 12.2 after the last treatment, respectively. The change over the course of the treatment was 17.5 ± 10.2 in MADRS and 13.8 ± 10.2 in BDI. The Pearson correlation coefficient between MADRS and BDI before and after the treatment was *r* = 0.46 and *r* = 0.62 respectively. The Pearson correlation coefficient of the change in MADRS and BDI over the course of the treatment was *r* = 0.50.

**Figure 3:**
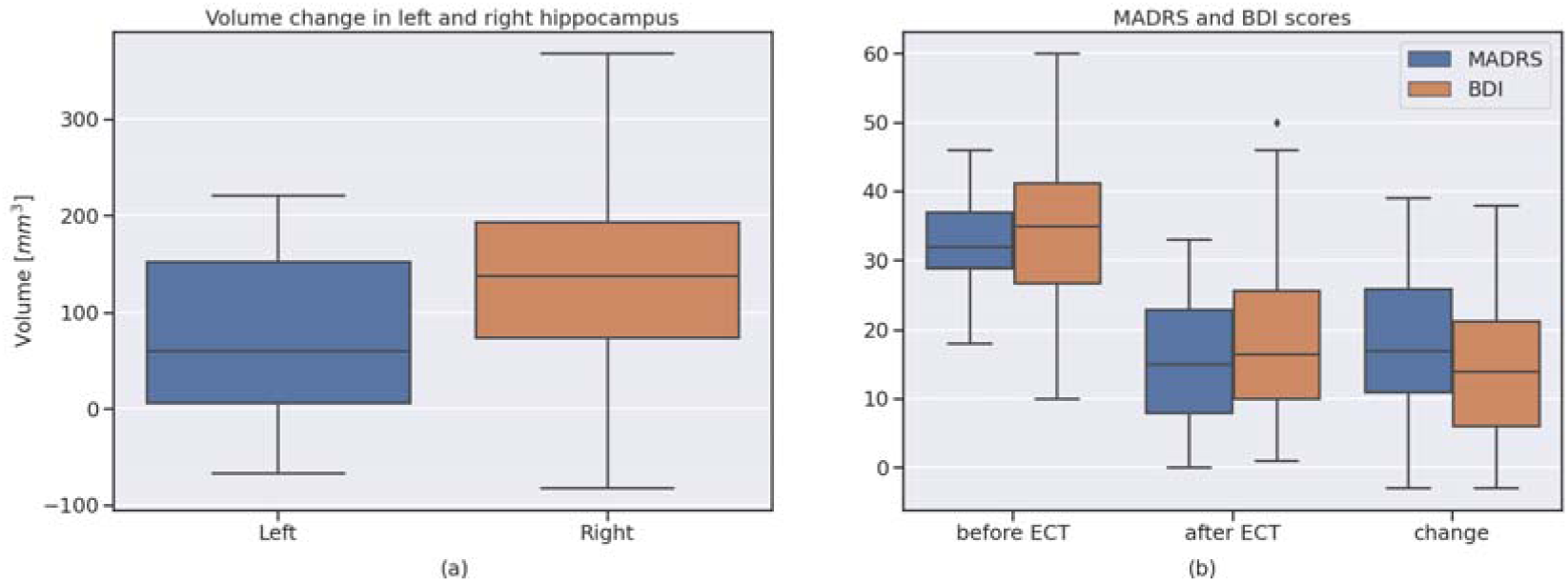
Volume change and clinical outcome: (a) Volume change in left and right hippocampus. (b) Montgommery and Åsberg depression rating scale (MADRS) and Beck depression inventory (BDI) scores before and after the electroconvulsive therapy (ECT) treatment, and the change after the treatment.

### Exploring EEG Models for Volume Change

Tables 1 and 2 show the *t*-values associated with each parameter in the successively simplified models for the volume change in the hippocampus in the right and left hemispheres, respectively. The tables also show *R*^2^, the *p*-value, and AIC for the successively simplified models. The regressor with the lowest associated *t*-value is marked in red for each model, and the best model is marked in green. The best model for the volume change in the right hippocampus has 4 parameters: the number of sessions, PS, and the energy in the alpha and beta bands, *F*(4, 24) = 5.85, *p* = 0.002, *R*^2^ = 0.49. The best model for the volume change of the hippocampus in the left hemisphere (Table 2), has 3 parameters: the number of sessions, cumulative charge, and LU, *F*(3, 25) = 6.46 *R*^2^ = 0.44. The two models for volume change in both hemispheres are further detailed in the supplementary materials.

**Table 1:** Table of t-values of the successively simplified models of volume change in the right hippocampus. Each column lists the t-values of the regressors in a particular linear model. The t-value with the lowest magnitude in each model is marked in red and is not included in the next model. The left-most column lists the name of each regressor. The lowest three rows show the, p-value, and AIC of each model. The model minimizing AIC is marked in green, and shown in detail in the supplementary materials.

| Name of regressor | Number of regressors |  |  |  |  |  |  |  |  |  |  |  |  |  |
| --- | --- | --- | --- | --- | --- | --- | --- | --- | --- | --- | --- | --- | --- | --- |
|  | 14 | 13 | 12 | 11 | 10 | 9 | 8 | 7 | 6 | 5 | 4 | 3 | 2 | 1 |
| Number of sessions | 1.54 | 1.60 | 1.66 | 1.66 | 1.78 | 1.80 | 1.53 | 1.29 | 1.31 | 2.21 | 4.27 | 3.18 | 2.84 | 3.14 |
| Cumulative charge | -0.819 | -0.87 | -0.94 | -1.00 | -1.20 | -1.18 | -0.83 |  |  |  |  |  |  |  |
| Cumulative seizure duration | 0.727 | 0.92 | 0.95 | 1.23 | 1.14 | 1.17 | 1.30 | 1.25 | 1.24 | 0.66 |  |  |  |  |
| Age | 0.935 | 0.97 | 1.02 | 1.05 | 1.18 | 1.26 | 0.89 | 0.35 |  |  |  |  |  |  |
| Sex | -0.06 |  |  |  |  |  |  |  |  |  |  |  |  |  |
| PS | -0.80 | -1.12 | -1.18 | -1.54 | -1.45 | -1.45 | -1.93 | -2.90 | -2.94 | -2.68 | 2.82 | -1.54 |  |  |
| Max sustained coherence | 0.32 | 0.37 | 0.39 |  |  |  |  |  |  |  |  |  |  |  |
| Time to peak coherence | -0.75 | -0.79 | -0.82 | -0.82 | 1.12 | -1.09 | -1.23 | -1.16 | -1.22 |  |  |  |  |  |
| Power law slope (lsq, R) | -0.06 | -0.06 |  |  |  |  |  |  |  |  |  |  |  |  |
| Power law slope (lu, R) | -0.09 | -0.10 | -0.83 | -0.77 | -1.02 | -0.99 |  |  |  |  |  |  |  |  |
| Energy delta (fraction, R) | -0.27 | -0.37 | -0.39 | -0.76 |  |  |  |  |  |  |  |  |  |  |
| Energy theta (fraction, R) | -0.35 | -0.47 | -0.49 | -0.85 | -0.38 |  |  |  |  |  |  |  |  |  |
| Energy alpha (fraction, R) | -1.99 | -2.07 | -2.15 | -2.17 | -2.06 | -2.36 | -2.24 | -2.41 | -2.57 | -2.62 | -2.57 |  |  |  |
| Energy beta (fraction, R) | 2.49 | 2.61 | 2.70 | 2.75 | 3.06 | 3.16 | 3.09 | 3.18 | 3.36 | 3.23 | 3.21 | 1.75 | 0.97 |  |
| $R^2$ | 0.596 | 0.596 | 0.596 | 0.592 | 0.578 | 0.575 | 0.553 | 0.538 | 0.535 | 0.503 | 0.494 | 0.354 | 0.293 | 0.267 |
| p-value | 0.237 | 0.161 | 0.103 | 0.065 | 0.026 | 0.026 | 0.019 | 0.012 | 0.006 | 0.004 | 0.002 | 0.011 | 0.011 | 0.004 |
| AIC | -125.445 | -127.439 | -129.432 | -131.164 | -133.494 | -133.951 | -134.494 | -135.510 | -137.338 | -137.434 | -138.886 | -133.824 | -133.192 | -134.154 |

**Table 2:**
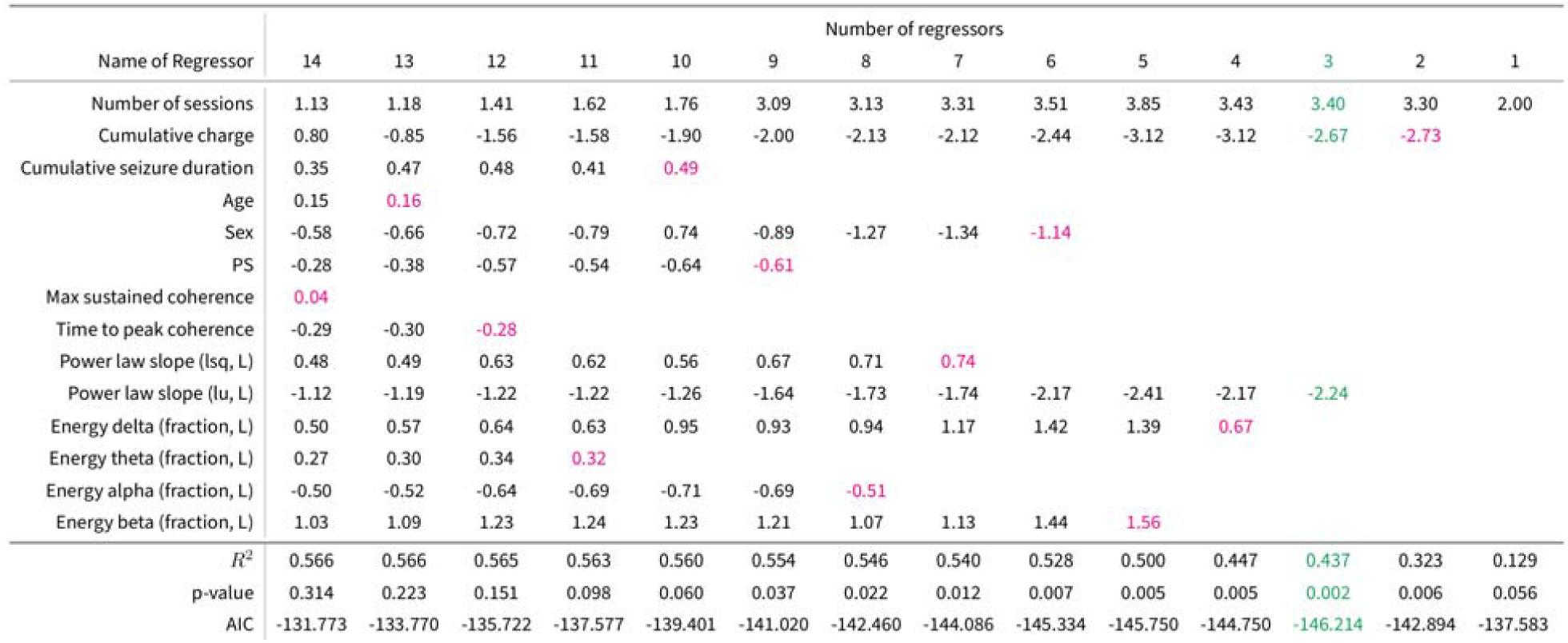
Table of *t*-values of the successively simplified models of volume change in the left hippocampus. Each column lists the *t*-values of the regressors in a particular linear model. The *t*-value with the lowest magnitude in each model is marked in red and is not included in the next model. The left-most column lists the name of each regressor. The lowest three rows show the, *p*-value, and AIC of each model. The model minimizing AIC is marked in green, and shown in detail in the supplementary materials.

### Exploring models for clinical outcome

The best model for change in MADRS, marked in green in Table 3, has 3 parameters: cumulative charge, age, and TPC, *F*(3, 25) = 4.28, *p* = 0.014, *=* 0.34. The model minimizing AIC for BDI in Table 4 has 8 parameters: volume change in the right hippocampus, cumulative charge, total seizure duration, age, sex, MSC, and least squares approximation of the power law, *F*(8, 17) = 4.61, *p* = 0.004 *=* 0.68. All four models minimize both AIC and cAIC.

**Table 3:**
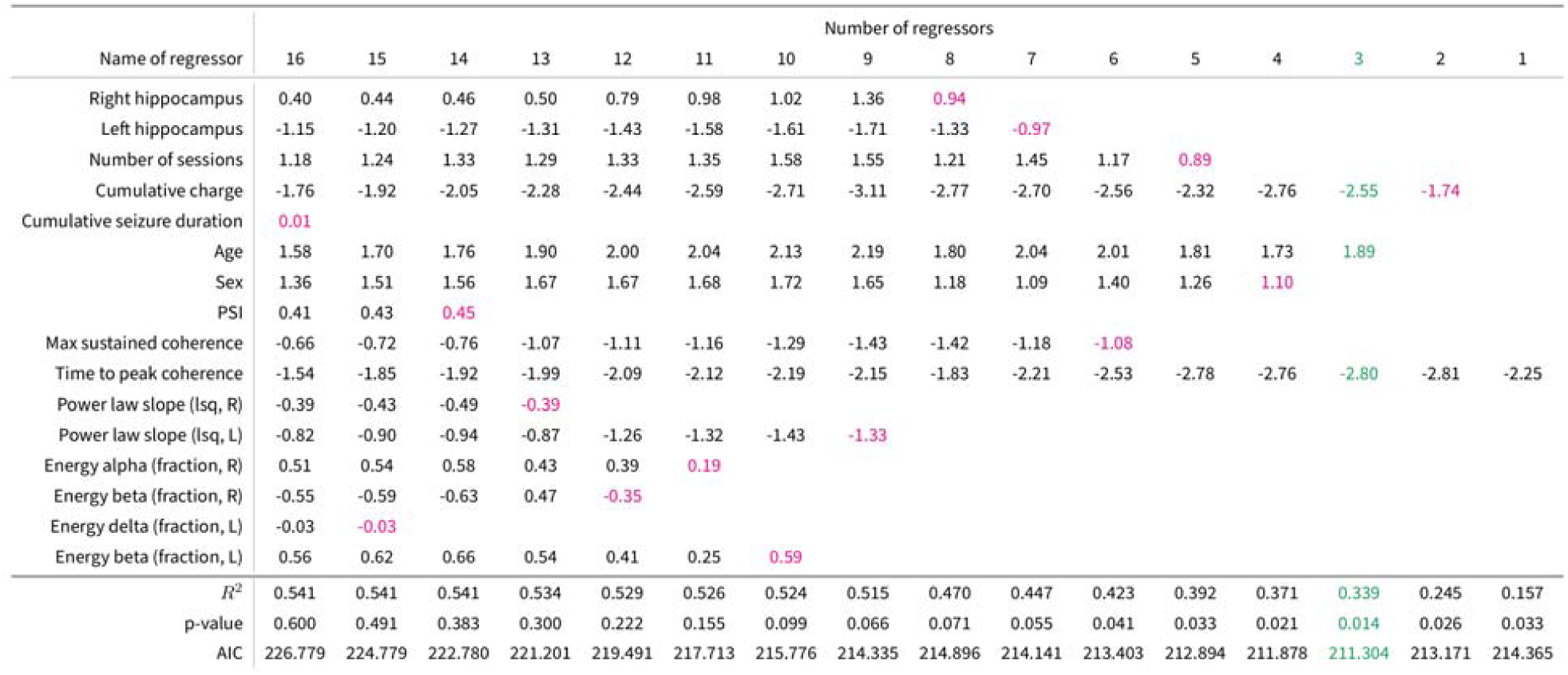
Table of *t*-values of the successively simplified models of change in MADRS score over the course of the treatment. Each column lists the t-values of the regressors in a particular linear model. The *t*-value with the lowest magnitude in each model is marked in red and is not included in the next model. The left-most column lists the name of each regressor. The lowest three rows show the, *p*-value, and AIC of each model. The model minimizing AIC is marked in green, and shown in detail in the supplementary materials.

**Table 4:** Table of *t*-values of the successively simplified models of change in BDI score over the course of the treatment. Each column lists the *t*-values of the regressors in a particular linear model. The *t*-value with the lowest magnitude in each model is marked in red and is not included in the next model. The left-most column lists the name of each regressor. The lowest three rows show the, *p*-value, and AIC of each model. The model minimizing AIC is marked in green, and shown in detail in the supplementary materials.

| Name of regressor | Number of regressors |  |  |  |  |  |  |  |  |  |  |  |  |  |  |  |
| --- | --- | --- | --- | --- | --- | --- | --- | --- | --- | --- | --- | --- | --- | --- | --- | --- |
|  | 16 | 15 | 14 | 13 | 12 | 11 | 10 | 9 | 8 | 7 | 6 | 5 | 4 | 3 | 2 | 1 |
| Right hippocampus | -0.81 | -1.02 | -1.11 | -1.41 | -1.30 | -1.60 | -1.58 | -1.65 | -1.63 | -1.90 |  |  |  |  |  |  |
| Left hippocampus | 0.08 |  |  |  |  |  |  |  |  |  |  |  |  |  |  |  |
| Number of sessions | 0.44 | 0.50 | 0.75 | 0.79 | 0.50 | 0.45 |  |  |  |  |  |  |  |  |  |  |
| Cumulative charge | -1.40 | -1.61 | -1.84 | -1.90 | -2.34 | -2.58 | -3.11 | -3.54 | -3.70 | -3.88 | -3.57 | -2.79 | -2.50 | -2.39 |  |  |
| Cumulative seizure duration | 1.43 | 1.51 | 1.60 | 1.86 | 1.81 | 1.94 | 2.66 | 2.67 | 2.65 | 2.79 | 2.09 |  |  |  |  |  |
| Age | 1.35 | 1.48 | 1.73 | 1.77 | 1.91 | 1.98 | 2.25 | 2.28 | 2.23 | 2.76 | 2.38 | 1.20 |  |  |  |  |
| Sex | 2.50 | 2.77 | 2.96 | 3.03 | 3.04 | 3.36 | 3.53 | 3.84 | 3.88 | 3.77 | 3.84 | 3.06 | 3.19 | 2.65 | 1.77 |  |
| PSI | 0.26 | 0.34 | 0.68 | 0.66 | 0.37 | 0.66 | 0.53 |  |  |  |  |  |  |  |  |  |
| Max sustained coherence | -0.28 | -0.30 |  |  |  |  |  |  |  |  |  |  |  |  |  |  |
| Time to peak coherence | -3.17 | -3.49 | -3.66 | -3.75 | -3.81 | -4.12 | -4.66 | -4.78 | -4.92 | -5.10 | -4.44 | -3.63 | -3.66 | -3.28 | -2.40 | -2.59 |
| Power law slope (lsq, R) | -1.55 | -1.68 | -1.85 | -2.53 | -2.70 | -2.93 | -3.15 | -3.24 | -3.26 | -2.92 | -2.78 | -2.37 | -2.10 |  |  |  |
| Power law slope (lsq, L) | -0.27 | -0.31 | -0.47 |  |  |  |  |  |  |  |  |  |  |  |  |  |
| Energy alpha (fraction, R) | 0.38 | 0.47 | 0.54 | 0.68 | 0.26 | 0.58 | 0.60 | 0.65 |  |  |  |  |  |  |  |  |
| Energy beta (fraction, R) | -0.30 | -0.34 | -0.47 | -0.68 |  |  |  |  |  |  |  |  |  |  |  |  |
| Energy delta (fraction, L) | 0.99 | 1.19 | 1.37 | 1.68 | 1.61 | 1.65 | 1.67 | 1.66 | 1.62 |  |  |  |  |  |  |  |
| Energy beta (fraction, L) | 0.34 | 0.37 | 0.50 | 0.68 | 0.26 |  |  |  |  |  |  |  |  |  |  |  |
| $R^2$ | 0.782 | 0.782 | 0.780 | 0.776 | 0.768 | 0.767 | 0.764 | 0.759 | 0.753 | 0.715 | 0.658 | 0.579 | 0.549 | 0.454 | 0.312 | 0.219 |
| $p$-value | 0.143 | 0.084 | 0.047 | 0.015 | 0.015 | 0.007 | 0.003 | 0.001 | 0.001 | 0.001 | 0.001 | 0.002 | 0.002 | 0.004 | 0.014 | 0.016 |
| AIC | 191.697 | 189.715 | 186.459 | 185.449 | 185.318 | 183.449 | 181.824 | 180.311 | 178.987 | 180.700 | 183.443 | 186.814 | 186.624 | 189.582 | 193.589 | 194.902 |

## Discussion

In this paper, we have introduced a novel EEG metric for assessment of ECT-induced seizures, the slope of the power-law in the EEG. We show that it is characteristic of seizures, and that it varies among patients. The slope of the power-law is estimated to be -3.80 and -3.61 in the left hemisphere using LSQ and LU respectively. In the right hemisphere, the slope was estimated to be -4.10 and -4.06 using LSQ and LU respectively. Furthermore, the power-law is complementary to common EEG measures, as demonstrated in linear models explaining the variation in volume change in the left hippocampus and change in clinical outcome, as measured by BDI.

The PSD is reasonably consistent between the patients, with two consistent peaks in the delta and beta bands, and most of the difference between patients is in the magnitude, see Fig 2b. It broadly follows a power-law distribution that we have characterized with two different methods, LSQ and LU. LSQ is sensitive to the power distribution over the whole frequency range from 3 Hz to 100 Hz, while LU is determined by the two points where the power-law intersects the PSD. These points are typically the peak in the beta band and the tail in the gamma band, or the peak in the delta band. The two measures are highly correlated in both hemispheres, but the Pearson correlation coefficient is higher in the right hemisphere. The high correlation coefficient indicates that the slope of the power law, *a*, is in this sense not sensitive to the choice of method used to estimate the slope. There is marked lateralization in the slope of the power-law, with the steepest slope seen in the hemisphere of electrode placement (RUL). Furthermore, the standard deviation is lower in the right than in the left hemisphere for both LSQ and LU, possibly reflecting that a stronger and more consistent electrical field in the right hemisphere has an impact on the seizure. The identified values for the slope of the power law during an ECT-induced seizure (∼ -3.6 - 4.1) stand in contrast to the slope of the power law reported under normal brain activity. Under normal brain activity Milstein et al. [14] found a = - 2 for frequencies less than 400 Hz and Miller et al. [15] found a = -2 for frequencies less than 75 Hz and a = -4 for frequencies between 80 and 500 Hz.

To our knowledge, no other study has investigated the relationship between volume changes in the hippocampus and ictal EEG parameters, which here are the slope of the power-law, PS, TPC, MSC and the energies in the delta, theta, alpha and beta bands. We systematically explored linear models relating the volume changes in the hippocampus to these EEG parameters. Our procedure of successively simplifying linear models yielded an optimal model, in terms of minimizing AIC, and also a series of significant models with fewer and more regressors in both hemispheres.

The number of ECT sessions appears in both best models for volume change in the hippocampus (right and left), and the cumulative charge appears in the best model for the left hippocampus, see Table 2. Despite the connection between the number of sessions, and the cumulative charge and seizure duration, the Pearson correlation coefficient between the number of sessions and cumulative charge is *r* = 0.51, and *r* = 0.79 between the number of sessions and cumulative seizure duration. This suggests that the gradual adjustment of stimulus dosage between treatments is not reflected in the seizure duration. The LU approximation of the slope of the power-law appears in the best model for the volume change in the left hippocampus.

We performed a similar explorative analysis of the relationship between EEG metrics and the volume changes in the hippocampus on one hand and clinical outcome as measured by MADRS or BDI on the other. The best model, according to AIC, for MADRS had 3 regressors: cumulative charge, age, and TPC, and 8 for BDI: volume change in the right hippocampus, cumulative charge, cumulative seizure duration, age, sex, TPC, and the energy in the delta frequency band. Cumulative charge, age and TPC appear in both models. The volume change in the left hippocampus does not appear in either the model for change in MADRS or BDI. However, the volume change in the right hippocampus appears in the model for change in BDI together with 7 other regressors, including cumulative charge and the slope of the power-law in the right hemisphere. Cumulative charge also appears in the model for the volume change in the left hippocampus.

There are conflicting reports as regards to the relationship between volume change in the hippocampus and ECT clinical outcome. Significant relationships have been found in a minority of studies [37, 38] although most studies could not document an association, as reflected by both mega-analysis [19] and meta-analyses [39, 40], see also review by Ousdal et al. [41]. The naturalistic study design of most studies investigating ECT-associated brain changes [19] complicates the interpretation of a potential relationship between volume change in the hippocampus and clinical outcome. The volume change in the hippocampus, at least in the right hemisphere correlates with the number of sessions, at the same time as ECT non-responders had more ECT sessions [19].

Estimates of the prevalence of depression, of any severity, using self-reported and clinician-rated scales are similar [42]. The agreement between the two types of rating scales is greater at follow-up than during the acute phase [43]. While the two scales give similar results in terms of the presence of symptoms, Uher et al. suggested that self-reported measures are preferred if not both are available [44].

The regression models of volume change outlined in this paper explain 49.4% of the variation in volume change in the right hippocampus and 43.7 % in the left hippocampus, see the *R*^2^ values in Tables 1 and 2. However, this must be weighed against the number of patients in this study (N = 29) and the number of regressors in the models. A similar critique holds against the models for MADRS and BDI. Choosing a model selection criterion, other than AIC or cAIC, which places a greater penalty on the number of regressors, could accommodate this critique. Model selection of this kind should be validated in prospective studies, as in the two studies by Kranaster et al. [45]. AIC balances between the goodness of fit of a model and its complexity, guiding the choice between overfitting and underfitting [35]. The power-law exponent from the EEG may be useful as a single-parameter characterization of the ECT-induced seizure, also because it shows seizure asymmetry between the hemispheres. The power-law characterization and the proposed models need further validation due to our relatively small sample size.

A previous study has sought to explain the volume change in terms of the electrical field induced by the stimulating electrodes [21]. The lateralization of the electric field is mirrored in the magnitude of the volume changes in the hippocampus. The volume changes are greater on the same side as the electrodes. Our model of volume change in the left hippocampus includes cumulative charge, indicating that the ECT stimulus dosage is important, not just the induced electrical field magnitude. The study by Takamiya et al. [22] found a correlation between volume change in the medial temporal lobe and the cumulative seizure duration, suggesting the seizure has some influence on the volume changes. However, the cumulative seizure duration did not figure in any of our models for volume change in the hippocampus.

Our finding of a lateralized power-law slope suggests a possible relation to ECT electrode placement and seizure quality, hence the measure could inform treatment decisions although further studies are necessary to investigate this possibility. Two previous studies have investigated the association between Thymatron IV EEG-measures and electrode position. Rasmussen et al. found a significant association between several parameters and Electrode positions [46], but Moss and Vaidya did not [47].

The lateralization is only seen in the slope of the power-law, and the energy in the theta band, but not the other frequency bands. Previous studies have found increased interhemispheric coherence to be a predictor of positive clinical outcome [9, 27, 48], and bilateral ECT leads to a higher degree of coherence in the slow frequency band (delta). It is unclear how interhemispheric coherence relates to the difference in power-law slope in the two hemispheres. Studies using other forms of seizure induction could help in disentangling the role of the seizure from that of the electrical stimulus or electrical field for the observed gray matter volumetric expansion seen with ECT. Since, Magnetic seizure therapy (MST, See Cretaz et al. for a review of the use of MST [49]) can deliver more focal stimulus than ECT, studies comparing the two treatment forms might also be used to probe the relationship between the lateralization of the induced seizure and the grey matter volume changes.

## Conclusion

The power-law in the PSD of the ictal EEG is characteristic of seizures, with some variation between patients. The slope of the power law was lateralized with larger absolute value seen in the right hemisphere suggesting an effect of the electrode placement. The power-law complements common EEG measures in explaining the variation in volume change in the hippocampus and change in BDI following ECT.

## Data Availability

All data produced in the present study are available upon reasonable request to the authors and written consent from study participants.

## CRediT author statemet

**Jakob Schreiner**: Conceptualization, Data curation, Formal analysis, Investigation, Project administration, Methodology, Software, Validation, Visualization, Writing – original draft

**Ute Kessler**: Conceptualization, Methodology, Validation, Writing – original draft – review and editing

**Ketil Oedegaard**: Validation, Writing – review and editing

**Kent Andre Mardal**: Conceptualization, Methodology, Supervision, Validation, Writing – original draft – review and editing

**Leif Oltedal**: Conceptualization, Methodology, Supervision, Validation, Writing – original draft – review and editing

## Abbreviations not common to the field

(PSD): Power spectral density
(TPC): Time to peak coherence
(MSC): Maximum sustained coherence
(LSQ): Least squares
(LU): Least upper,

## Acknowledgments

Thanks to Leila Marie Frid for help with accessing data, Ingrid Mossige for providing Freesurfer data processing, Lars Willas Dreyer and Bastian Zapf for help digitizing EEGs. We are grateful to Tom Eichele for advice, careful review and helpful comments on the manuscript. We are also grateful to all study participants.

