## Appendix A for "Exploring new EEG-parameters in electroconvulsive therapy"

### Appendix A – Digitizing EEGs From Paper Strips

The EEG paper strips generated by Thymatron IV were digitized by *in-house* developed software, which relied on image processing algorithms implemented in the computer vision library (open-cv). The complete algorithms are detailed in Appendix A and available at (https://github.com/expertanalytics/digeeg). Each EEG was carefully reviewed by JS, and segments exhibiting movement and muscle-related artifacts were discarded as recommended [1], see e.g. [2, 3].

The main strategy behind the digitization process is to reorient and rescale the images so they can be treated as coordinate systems, and then to extract the voltage traces and describe each one by an array of coordinates (time, voltage). The whole process can be described in five broad points:

- Each scan is split into smaller parts and resampled into the same size and orientation. This accounts for twisting of the paper during the scanning process. The splitting and resampling both rely on the black square fiduciary markers printed on the EEG paper-strips.
- Successive blurring and threshold operations remove the background millimeter pattern. If the black line denoting the duration of the seizure intersects the voltage traces, it is removed by a convolution with a rectangular kernel.
- The voltage traces are segmented using the contour finding tool in *open-cv*. The contours are filtered based on aspect ratio to distinguish them from the text and numbers and other artifacts. The segmented lines are still images.
- The voltage traces are converted into (time, voltage) pairs by treating the x-axis of the image as time, and y-axis as voltage.
- The final step is for the user to join the voltage traces together from the smaller parts created in step 1. Furthermore, the traces might not be segmented continuously due to smudges on the paper, poor quality scans or intersections with the line marking the seizure duration. This step also served as quality control. The traces are centered on zero by subtracting the mean of the voltage.
