## Supplementary materials 1 for "Exploring new EEG-parameters in electroconvulsive therapy"

### Right Hippocampus

```
Residuals:
      Min        1Q    Median        3Q       Max
-0.034810 -0.013976 -0.004786  0.010229  0.044714

Coefficients:
                            Estimate Std. Error t value Pr(>|t|)
(Intercept)                 0.056127   0.021076   2.663 0.013608 *
Number of Sessions          0.004417   0.001035   4.268 0.000267 ***
PS                         -0.043510   0.015449  -2.816 0.009559 **
Energy alpha (fraction, R) -0.812435   0.315832  -2.572 0.016714 *
Energy beta (fraction, R)   0.956760   0.297893   3.212 0.003733 **
---
Signif. codes:  0 '***' 0.001 '**' 0.01 '*' 0.05 '.' 0.1 ' ' 1

Residual standard error: 0.01973 on 24 degrees of freedom
Multiple R-squared:  0.4939,    Adjusted R-squared:  0.4095
F-statistic: 5.854 on 4 and 24 DF,  p-value: 0.001958
```

### Left Hippocampus

```
Residuals:
      Min        1Q    Median        3Q       Max
-0.031620 -0.011874  0.004686  0.015223  0.021138

Coefficients:
                            Estimate Std. Error t value Pr(>|t|)
(Intercept)                -6.603e-02  3.223e-02  -2.049  0.05113 .
Number of sessions          3.172e-03  9.319e-04   3.403  0.00225 **
Cumulative charge          -6.726e-06  2.519e-06  -2.670  0.01315 *
Power law slope (lu, left) -1.922e-02  8.566e-03  -2.244  0.03396 *
---
Signif. codes:  0 '***' 0.001 '**' 0.01 '*' 0.05 '.' 0.1 ' ' 1

Residual standard error: 0.01763 on 25 degrees of freedom
Multiple R-squared:  0.4365,    Adjusted R-squared:  0.3689
F-statistic: 6.455 on 3 and 25 DF,  p-value: 0.002185
```

### MADRS

```
Residuals:
    Min      1Q  Median      3Q     Max
-15.392  -4.134  -0.273   4.735  19.436
```

```
Coefficients:
                        Estimate Std. Error t value Pr(>|t|)
(Intercept)            35.767167   9.495674   3.767  0.00090 ***
Cumulative charge      -0.003329   0.001305  -2.551  0.01724 *
Age                     0.230515   0.122083   1.888  0.07066 .
Time to peak coherence -0.854310   0.305645  -2.795  0.00982 **
---
Signif. codes:  0 '***' 0.001 '**' 0.01 '*' 0.05 '.' 0.1 ' ' 1

Residual standard error: 8.382 on 25 degrees of freedom
Multiple R-squared:  0.3394,    Adjusted R-squared:  0.2602
F-statistic: 4.282 on 3 and 25 DF,  p-value: 0.01434
```

### BDI

```
Residuals:
    Min      1Q  Median      3Q     Max
-9.1283 -3.3608 -0.1561  3.0064 18.3357

Coefficients:
                             Estimate Std. Error t value Pr(>|t|)
(Intercept)                 -3.723011  19.106063  -0.195 0.847812
Right hippocampus          -64.786164  73.343997  -0.883 0.389385
Cumulative charge           -0.006338   0.001845  -3.434 0.003163 **
Cumulative seizure duration  0.019257   0.009473   2.033 0.057998 .
Age                          0.317608   0.166013   1.913 0.072724 .
Sex                         13.328216   3.693170   3.609 0.002167 **
Time to peak coherence      -1.626952   0.350878  -4.637 0.000236 ***
Power law slope (lsq, L)    -9.905629   4.577279  -2.164 0.044985 *
Energy delta (fraction, L)   7.972486  14.187021   0.562 0.581480
---
Signif. codes:  0 '***' 0.001 '**' 0.01 '*' 0.05 '.' 0.1 ' ' 1

Residual standard error: 7.189 on 17 degrees of freedom
  (3 observations deleted due to missingness)
Multiple R-squared:  0.6847,    Adjusted R-squared:  0.5363
F-statistic: 4.614 on 8 and 17 DF,  p-value: 0.003905
```
